# International risk of the new variant COVID-19 importations originating in the United Kingdom

**DOI:** 10.1101/2021.01.09.21249384

**Authors:** Zhanwei Du, Lin Wang, Bingyi Yang, Sheikh Taslim Ali, Tim K. Tsang, Songwei Shan, Peng Wu, Eric H. Y. Lau, Benjamin J. Cowling, Lauren Ancel Meyers

**Affiliations:** WHO Collaborating Centre for Infectious Disease Epidemiology and Control, School of Public Health, LKS Faculty of Medicine, The University of Hong Kong, Hong Kong Special Administrative Region, China; Laboratory of Data Discovery for Health, Hong Kong Science and Technology Park, Hong Kong Special Administrative Region, China; University of Cambridge, Cambridge CB2 3EH, UK; The University of Texas at Austin, Austin, Texas 78712, The United States of America; Santa Fe Institute, Santa Fe, New Mexico, The United States of America

## Abstract

A fast-spreading SARS-CoV-2 variant identified in the United Kingdom in December 2020 has raised international alarm. We estimate that, in all 15 countries analyzed, there is at least a 50% chance the variant was imported by travelers from the United Kingdom by December 7th.

---

The United Kingdom (UK) has detected a new variant of SARS-CoV-2 from samples initially taken in Kent on September 20th and London on September 21st, 2020 (1), which was found associated with increased transmissibility (2). The UK government tightened measures in London and southeast England in mid-December to mitigate the transmission of the fast-spreading virus variant that includes deletions at amino acid sites 69 and 70 of the spike protein (3). On January 5th, 2021, England initiated a national lockdown including closure of all schools and non-essential businesses until mid-February (4). By December 20th, over 40 countries had implemented travel restrictions on travellers from the UK (5). The new variant was subsequently reported worldwide, including in the USA (6), Spain, Sweden and France, and might be spreading without detection in countries with limited virus sequencing capacity (5).

We collected the data from 15 countries and estimated the probability of introduction of this new variant by travellers from the UK to each of these countries and the extent of local transmission, based on the changing proportion of the new variant among infections identified in the UK (2) and population mobility from UK to each country, as estimated from Facebook Data for Good. Ireland had the highest importation risk between September 22 and December 7, 2020. By October 22 (a month after its first detection in the UK), 10 of the 15 countries had at least a 50% chance of receiving one imported case from the UK (**Figure 1**), except for Romania, Portugal, Cyprus, India, and United States, while they all had exceeded this risk threshold by November 1.

**Figure 1.**
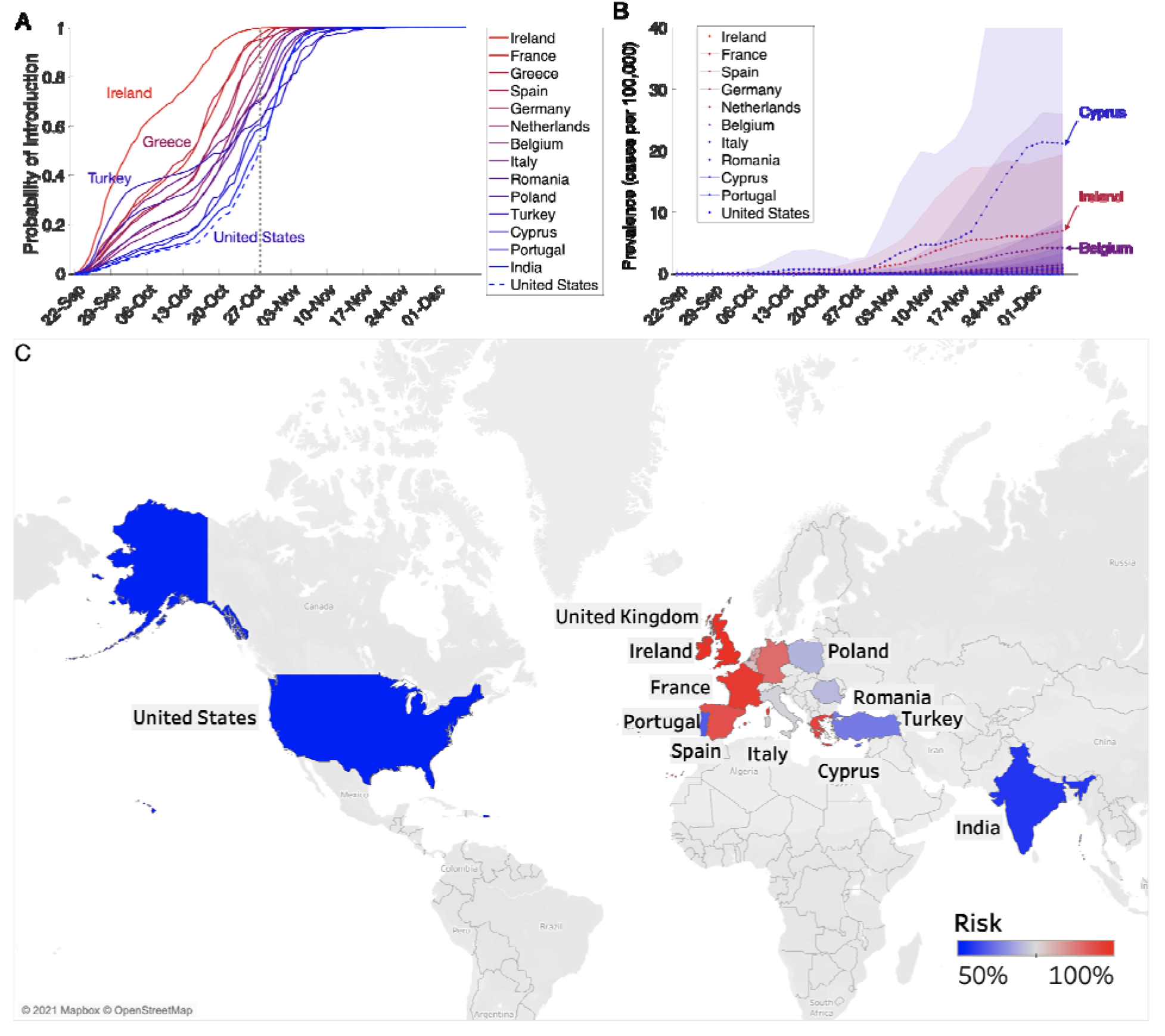
Estimated risks for introduction of the 501Y variant of SARS-CoV-2 from the UK to other 15 countries before December 7, 2020. (A) The probability that at least one person infected with the new COVID-19 variant has arrived at the target country from the UK by the date indicated on the x-axis, based on Facebook mobility data. The dotted gray vertical line indicates October 28, 2020, the date that the introduction risk for the USA surpasses 50%; line colors correspond to the relative risk of importations as of that date. (B) Estimated daily prevalence of the 501Y variant of SARS-CoV-2 in 11 countries between September 22 and December 7, 2020, assuming that the variant is σ= 50% more transmissible than the 501N variant (11). Points and bands indicate means and standard deviations based on 100 simulations. (C) Prosbability of at least one variant importation by October 28, 2020. Grey indicates countries/regions where mobility data were not available.

Using COVID-19 hospital admission data, we further estimated the local prevalence of the new virus variant in 11 of the 15 countries, assuming that the new variant is 50% more transmissible than the circulating 501N strain (**Figure 1**). The variant appears to have ascended fastest in Ireland before slowing in mid November and is expected to be spreading rapidly in many of the other countries. As of December 7, Cyprus has the highest expected prevalence of the variant (13 [95% CI: 0-79] cases per 100,000) and Ireland has the highest expected proportion of COVID-19 cases caused by the new variant (6% [95% CI: 0-38%]) (**Figures 1, S1 and S2**).

These projections suggested that countries with substantial population movement from the UK were likely to harbor cases of the new variant by late October, 2020. Our conclusions were based on several key assumptions. The mobility data, which includes ∼3 million trips from the UK to the 15 focal countries, might be demographically biased by the user profile of Facebook, a major social media company with ∼2.7 billion monthly active users in the third quarter of 2020 (7). We assume that all introductions during this early period occurred via asymptomatic travelers from the UK and ignore possible importations from other countries or by symptomatic cases traveling to seek healthcare. A sensitivity analysis suggests that these assumptions may cause a downward bias in the estimated rates of global expansion (Figure S3). Furthermore, we assume a 10-day lag between infection and hospitalization based on estimates from the United States (8) and Europe (9) and estimate the daily prevalence of the new strain using the method introduced in ref. (2), under the assumptions that the two variants (501Y and 501N) share the same natural history (2) and symptomatic proportion (10,11). Should future studies reveal significant epidemiological differences between the variant and wildtype, then these estimates can be readily updated using the full equations provided in ref. (2).

## Data Availability

Not applicable

## Acknowledgements

Financial support was provided by the Health and Medical Research Fund, Food and Health Bureau, Government of the Hong Kong Special Administrative Region (grant no. COVID190118), the US National Institutes of Health (grant no. R01 AI151176) and CDC COVID Supplement (grant no. U01IP001136).

Dr. Zhanwei Du is a research assistant professor in the School of Public Health, LKS Faculty of Medicine, The University of Hong Kong, Hong Kong Special Administrative Region, China. He develops mathematical models to elucidate the transmission dynamics, surveillance and control of infectious diseases.

## References

1. Mutant coronavirus in the United Kingdom sets off alarms, but its importance remains unclear [Internet]. 2020 [cited 2021 Jan 2]. Available from: https://www.sciencemag.org/news/2020/12/mutant-coronavirus-united-kingdom-sets-alarms-its-importance-remains-unclear

2. Leung K, Shum MHH, Leung GM, Lam TTY, Wu JT. Early transmissibility assessment of the N501Y mutant strains of SARS-CoV-2 in the United Kingdom, October to November 2020. Eurosurveillance. 2021 Jan 7;26(1):2002106.

3. Kupferschmidt K. Fast-spreading U.K. virus variant raises alarms. Science. 2021 Jan 1;371(6524):9–10.

4. Clark N. National lockdown – Boris Johnson orders nation to stay at home until middle of February as 13.5m Brits get C. The Sun [Internet]. 2021 Jan 5 [cited 2021 Jan 7]; Available from: https://www.thesun.co.uk/news/politics/13648237/national-lockdown-stay-home-boris-johnson-announcement/

5. BBC News. Coronavirus: Cases of new variant appear worldwide. BBC [Internet]. 2020 Dec 26 [cited 2021 Jan 2]; Available from: https://www.bbc.com/news/world-europe-55452262

6. CDC. Emerging SARS-CoV-2 Variants [Internet]. 2020 [cited 2021 Jan 2]. Available from: https://www.cdc.gov/coronavirus/2019-ncov/more/science-and-research/scientific-brief-emerging-variants.html

7. Number of monthly active Facebook users worldwide as of 3rd quarter 2020 [Internet]. [cited 2021 Jan 3]. Available from: https://www.statista.com/statistics/264810/number-of-monthly-active-facebook-users-worldwide/

8. Aleta A, Martín-Corral D, Pastore Y Piontti A, Ajelli M, Litvinova M, Chinazzi M, et al. Modelling the impact of testing, contact tracing and household quarantine on second waves of COVID-19. Nat Hum Behav [Internet]. 2020 Aug 5; Available from: http://dx.doi.org/10.1038/s41562-020-0931-9

9. ISARIC. International Severe Acute Respiratory and Emerging Infections Consortium (ISARIC). COVID-19 Report: 19 [Internet]. 2020 [cited 2021 Feb 1]. Available from: https://media.tghn.org/medialibrary/2020/05/ISARIC_Data_Platform_COVID-19_Report_19MAY20.pdf

10. King’s College London. No evidence of change in symptoms from new coronavirus variant [Internet]. King’s College London. 2021 [cited 2021 Mar 3]. Available from: https://www.kcl.ac.uk/news/no-evidence-change-symptoms-new-coronavirus-variant

11. Davies NG, Abbott S, Barnard RC, Jarvis CI, Kucharski AJ, Munday JD, et al. Estimated transmissibility and impact of SARS-CoV-2 lineage B.1.1.7 in England. Science [Internet]. 2021 Mar 3; Available from: http://dx.doi.org/10.1126/science.abg3055

