## Appendix for "International risk of the new variant COVID-19 importations originating in the United Kingdom"

**Affiliations:**

**Supplement**

**Table S1. Model Parameters and Data Sources.**

| **Symbol** | **Description** | **Values** | **Sources** |
| --- | --- | --- | --- |
| 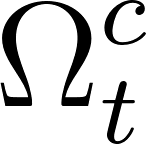<https://www.codecogs.com/eqnedit.php?latex=%5COmega%5E%7Bc%7D_%7Bt%7D#0> | Number of travelers from the UK to country *c* at time *t* | Daily mobility | Facebook Data for Good (1) |
| 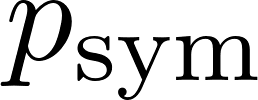<https://www.codecogs.com/eqnedit.php?latex=p_%7B%5Ctext%7Bsym%7D%7D#0> | Proportion of infections that are symptomatic | 57% | Ref. (2),  assuming 501Y and 501N have the same symptomatic proportion (3,4) |
| 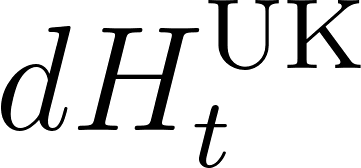<https://www.codecogs.com/eqnedit.php?latex=dH%5E%7B%5Ctext%7BUK%7D%7D_t#0> | Number of COVID-19 hospital admissions in the UK at time *t* | Daily admissions | Refs. (5,6) |
| 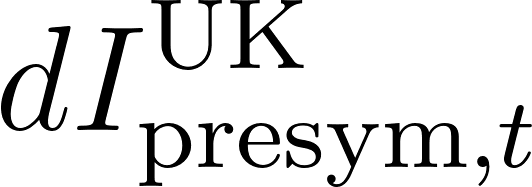<https://www.codecogs.com/eqnedit.php?latex=%20dI%5E%7B%5Ctext%7BUK%7D%7D_%7B%5Ctext%7Bpresym%7D%2Ct%7D%20#0> | Number of new pre-symptomatic variant infections in the UK at time *t* | Daily cases | Estimated |
| 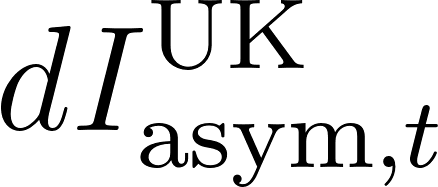<https://www.codecogs.com/eqnedit.php?latex=%20dI%5E%7B%5Ctext%7BUK%7D%7D_%7B%5Ctext%7Basym%7D%2Ct%7D%20#0> | Number of new asymptomatic variant infections in the UK at time *t* | Daily cases | Estimated |
| 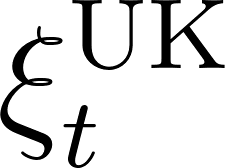<https://www.codecogs.com/eqnedit.php?latex=%5Cxi%5E%7B%5Ctext%7BUK%7D%7D_%7Bt%7D#0> | Prevalence of asymptomatic variant cases as a percentage of the UK population at time *t* | Daily prevalence | Estimated |
| 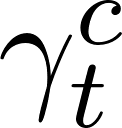<https://latex-staging.easygenerator.com/eqneditor/editor.php?latex=%5Cgamma_t%5Ec#0> | Rate of variant introductions from the UK to country *c* on day *t* | Daily rate | Estimated |
| 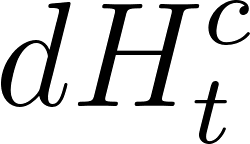<https://www.codecogs.com/eqnedit.php?latex=dH%5E%7Bc%7D_t#0> | Number of COVID-19 hospital admissions in country *c* at time *t* | Daily admissions | Refs. (5,6) |
| 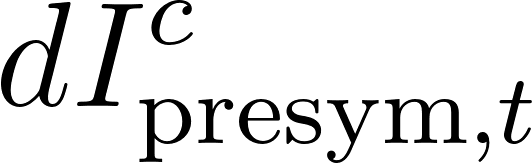<https://www.codecogs.com/eqnedit.php?latex=%20dI%5E%7Bc%7D_%7B%5Ctext%7Bpresym%7D%2Ct%7D%20#0> | Number of new pre-symptomatic variant infections in country *c* at time *t* | Daily cases | Estimated |
| 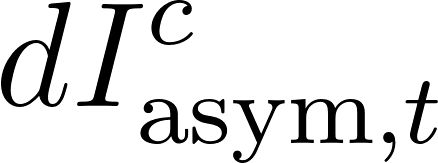<https://www.codecogs.com/eqnedit.php?latex=%20dI%5E%7Bc%7D_%7B%5Ctext%7Basym%7D%2Ct%7D%20#0> | Number of new asymptomatic variant infections in country *c* at time *t* | Daily cases | Estimated |
| 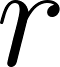<https://www.codecogs.com/eqnedit.php?latex=r#0> | Proportion of symptomatic cases that are hospitalized | 3.4/77 | Ref. (7) |
| 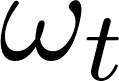<https://www.codecogs.com/eqnedit.php?latex=%5Comega_t#0> | Proportion of the 501Y variant (with deletions at amino acid sites 69/70 of the S protein) among new SARS-CoV-2 cases at time *t* | Proportion of sequenced SARS-CoV-2 specimens in UK (weekly data) | Ref. (8) |
| 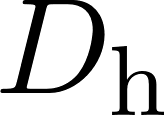<https://www.codecogs.com/eqnedit.php?latex=D_%7B%5Ctext%7Bh%7D%7D#0> | Expected delay from infection to hospital admission | 10 days | 5 days from infection to symptom onset (9); 5 days from symptom onset to hospital admission (10,11) |
| 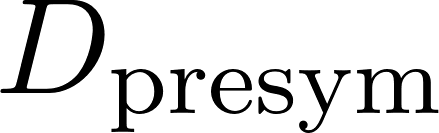<https://www.codecogs.com/eqnedit.php?latex=D_%5Ctext%7Bpresym%7D#0> | Expected incubation period between infection and symptom onset for symptomatic cases | 5 days | Ref. (9) |
| 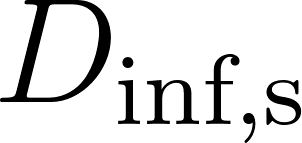<https://www.codecogs.com/eqnedit.php?latex=D_%5Ctext%7Binf%2Cs%7D#0> | Expected time from infection to recovery, for symptomatic cases | 11 days | Sum of expected 5-day incubation (9) and expected 6-day symptomatic (10)  period |
| 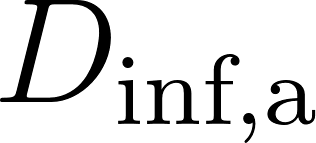<https://www.codecogs.com/eqnedit.php?latex=D_%5Ctext%7Binf%2Ca%7D#0> | Expected time from infection to recovery, for asymptomatic cases | 11 days | Ref. (10) |
| 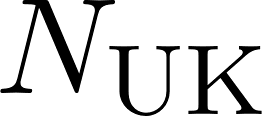<https://www.codecogs.com/eqnedit.php?latex=N_%7B%5Ctext%7BUK%7D%7D#0> | Population of United Kingdom | 66,796,807 (2019) | Ref. (12) |
| 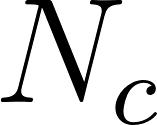<https://www.codecogs.com/eqnedit.php?latex=N_%7Bc%7D#0> | Population of country *c* | 2019 populations | Ref. (13) |
| 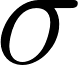<https://www.codecogs.com/eqnedit.php?latex=%5Csigma#0> | Ratio of the reproduction numbers for the 501Y versus 501N variant | 1.25, 1.5, and 1.75 | Assumed based on Refs. (14–16) |

### **Data**

To estimate the daily number of passengers traveling between the UK and other countries, we obtained daily mobility data from Facebook Data for Good (1). Based on geolocation data from Facebook mobile users that enable the ‘location history’ feature, Facebook constructed anonymized origin–destination mobility matrices between countries. However, Facebook only reports mobility flows from the UK to other countries when the daily number of recorded trips is at least 1000. We excluded countries from our analysis that had over 30% days between September 22 and October 21, 2020 falling below this threshold. For the countries we analyzed, we assumed a random number of trips (uniformly distributed between 0 and 1000) on each day of missing data. The data for those countries include ~3 million trips originating in the UK between September 22 and December 14, 2020.

### **Methods**

***Risk of COVID-19 variant introduction via infected travelers from the UK***

To estimate the probability at least one case has been introduced from the UK into a given country, we first estimate the prevalence of pre-symptomatic and asymptomatic cases in the UK and then use mobility data to estimate the likelihood of introductions.

Our notation and parameter values are provided in **Table S1**. Briefly, we assume that a fraction
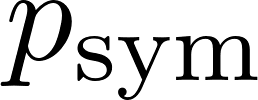
<https://www.codecogs.com/eqnedit.php?latex=p_%7B%5Ctext%7Bsym%7D%7D#0> of cases eventually develop symptoms and the remaining 1-
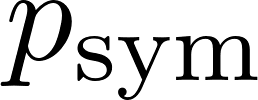
<https://www.codecogs.com/eqnedit.php?latex=p_%7B%5Ctext%7Bsym%7D%7D#0> remain asymptomatic throughout the course of their infection. The total periods of infection for asymptomatic and symptomatic cases are
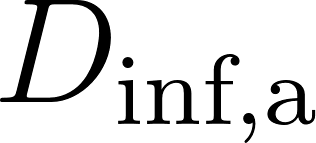
<https://www.codecogs.com/eqnedit.php?latex=D_%5Ctext%7Binf%2Ca%7D#0> and
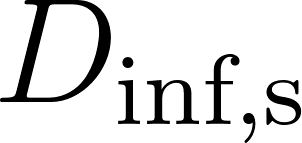
<https://www.codecogs.com/eqnedit.php?latex=D_%5Ctext%7Binf%2Cs%7D#0> days, respectively, with symptomatic cases developing symptoms following a
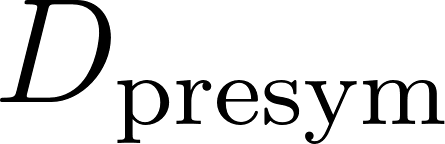
<https://www.codecogs.com/eqnedit.php?latex=D_%7B%5Ctext%7Bpresym%7D%7D#0> day incubation period. A proportion is denoted
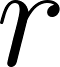
<https://www.codecogs.com/eqnedit.php?latex=r#0> of symptomatic cases require hospitalization and enter the hospital
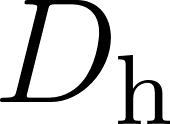
<https://www.codecogs.com/eqnedit.php?latex=D_%7B%5Ctext%7Bh%7D%7D#0> days following infection. We use
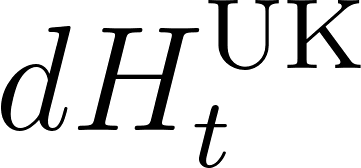
<https://www.codecogs.com/eqnedit.php?latex=dH%5E%7B%5Ctext%7BUK%7D%7D_t#0> and
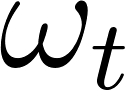
<https://www.codecogs.com/eqnedit.php?latex=%5Comega_t#0> to denote the number of new COVID-19 hospital admissions and the proportion of the new variant among all sequenced SARS-CoV-2 samples in the UK on day *t*, respectively. We assume that the numbers of new pre-symptomatic and asymptomatic variant-caused infections in the UK at time *t* are given by

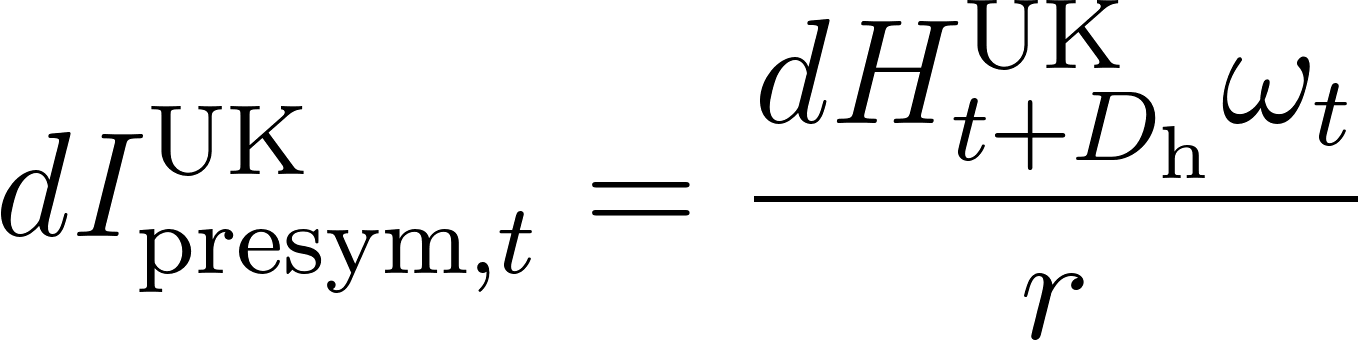
<https://www.codecogs.com/eqnedit.php?latex=dI%5E%7B%5Ctext%7BUK%7D%7D_%7B%7B%5Ctext%7Bpresym%7D%7D%2Ct%7D%3D%5Cfrac%7BdH%5E%7B%5Ctext%7BUK%7D%7D_%7Bt%2BD_%7B%5Ctext%20h%7D%7D%5Comega_t%7D%7Br%7D#0>

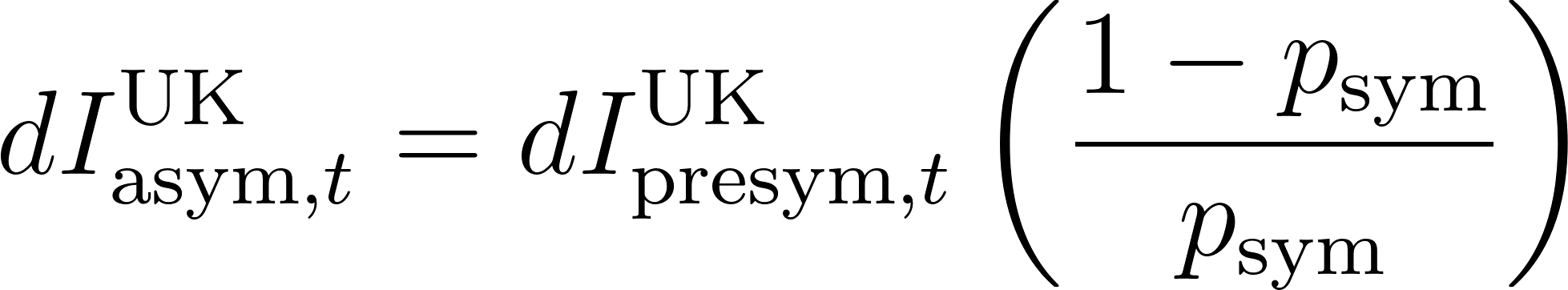
<https://www.codecogs.com/eqnedit.php?latex=dI%5E%7B%5Ctext%7BUK%7D%7D_%7B%5Ctext%7Basym%7D%2Ct%7D%20%3D%20dI%5E%7B%5Ctext%7BUK%7D%7D_%7B%5Ctext%7Bpresym%7D%2Ct%7D%5Cleft(%5Cfrac%7B1-%7Bp_%7B%5Ctext%7Bsym%7D%7D%7D%20%7D%7Bp_%7B%5Ctext%7Bsym%7D%7D%7D%5Cright)%20#0>.

Then, the numbers of pre-symptomatic and asymptomatic cases in the UK at time *t* are given by

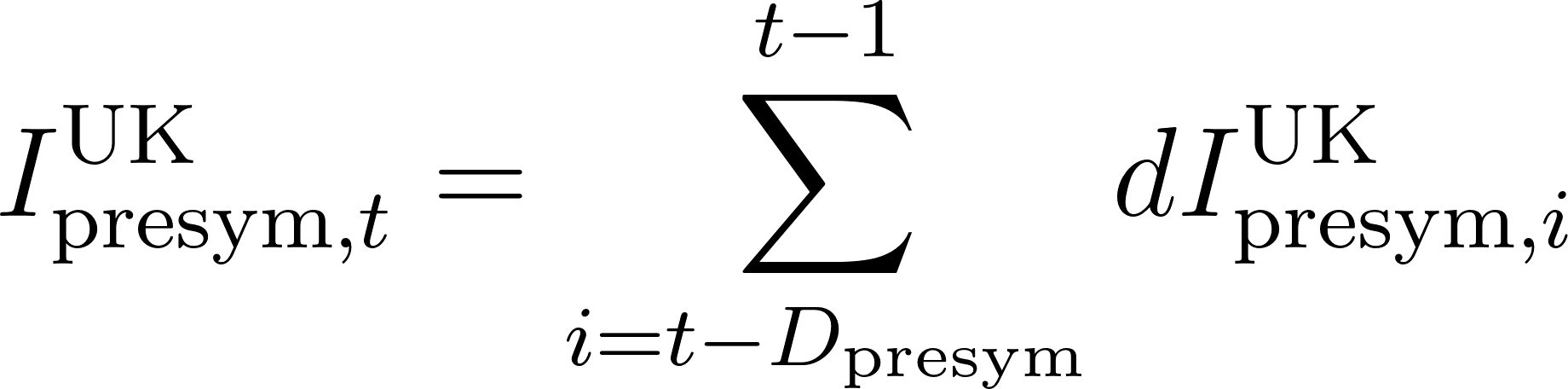
<https://www.codecogs.com/eqnedit.php?latex=%20I%5E%7B%5Ctext%7BUK%7D%7D_%7B%5Ctext%7Bpresym%7D%2C%20t%7D%20%3D%20%5Csum_%7Bi%20%3D%20t%20-%20D_%7B%5Ctext%7Bpresym%7D%7D%7D%5E%7Bt-1%7D%20dI%5E%7B%5Ctext%7BUK%7D%7D_%7B%5Ctext%7Bpresym%7D%2C%20i%7D%20%20#0>

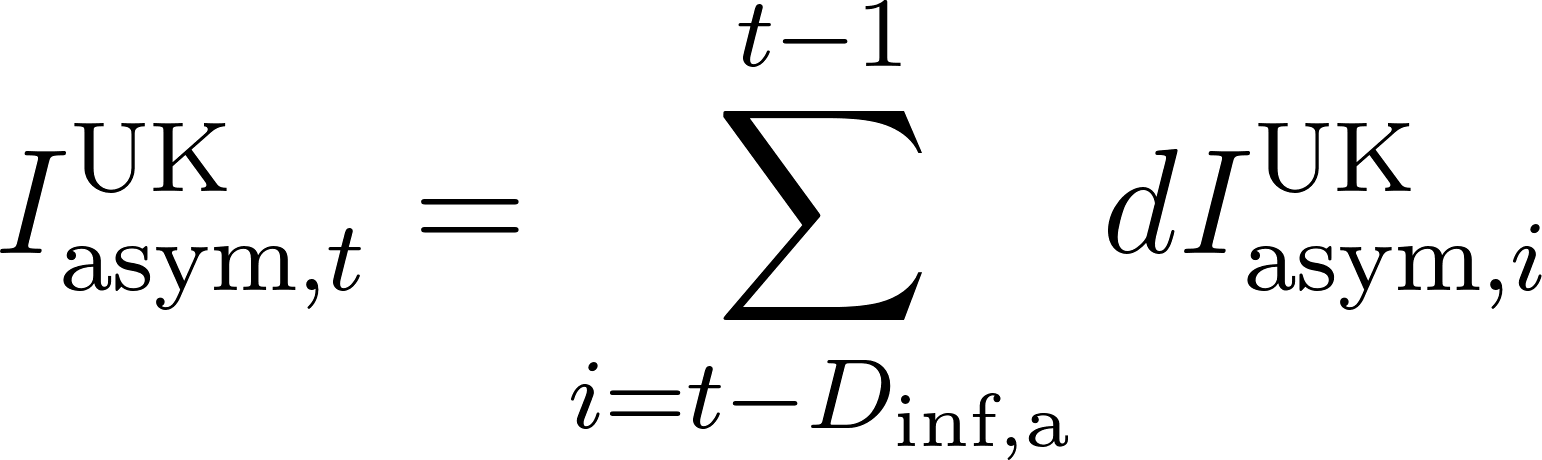
<https://www.codecogs.com/eqnedit.php?latex=%20I%5E%7B%5Ctext%7BUK%7D%7D_%7B%5Ctext%7Basym%7D%2Ct%7D%20%3D%20%5Csum_%7Bi%20%3D%20t%20-%20D_%7B%5Ctext%7Binf%2Ca%7D%7D%7D%5E%7Bt-1%7D%20dI%5E%7B%5Ctext%7BUK%7D%7D_%7B%5Ctext%7Basym%7D%2C%20i%7D%20#0>

The prevalence of asymptomatic cases infected by the variant as a proportion of the UK population is given by

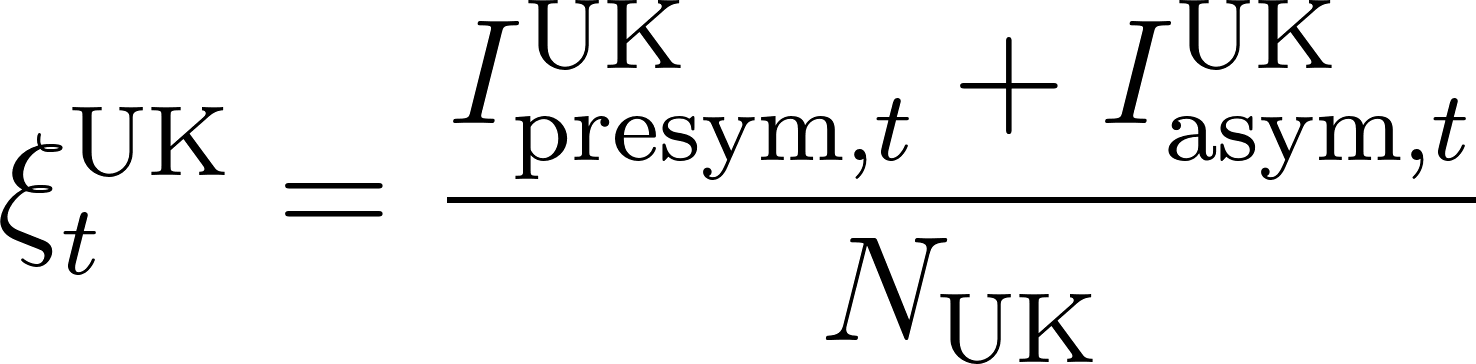
<https://www.codecogs.com/eqnedit.php?latex=%5Cxi%5E%7B%5Ctext%7BUK%7D%7D_%7Bt%7D%20%3D%20%5Cfrac%7B%20I%5E%7B%5Ctext%7BUK%7D%7D_%7B%5Ctext%7Bpresym%7D%2C%20t%7D%2B%20I%5E%7B%5Ctext%7BUK%7D%7D_%7B%5Ctext%7Basym%7D%2Ct%7D%7D%7BN_%7B%5Ctext%7BUK%7D%7D%7D%20#0>

where
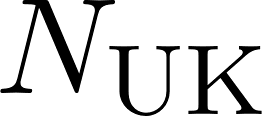
<https://www.codecogs.com/eqnedit.php?latex=N_%7B%5Ctext%7BUK%7D%7D#0> is the population size of the UK*.*

Assuming that travelers from the UK to other countries are infected with the variant according to the overall prevalence of pre-symptomatic and asymptomatic cases (assuming symptomatic cases will not travel), the rate of introductions from the UK to country *c* on day *t* is approximated by

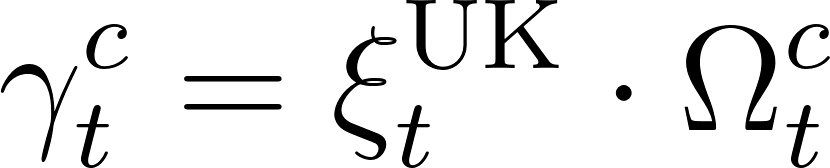
<https://www.codecogs.com/eqnedit.php?latex=%20%5Cgamma%5E%7Bc%7D_%7Bt%7D%3D%5Cxi%5E%7B%5Ctext%7BUK%7D%7D_%7Bt%7D%20%5Ccdot%20%5COmega%5E%7Bc%7D_%7Bt%7D%20#0>[,](https://www.codecogs.com/eqnedit.php?latex=%20%5Cgamma%5E%7Bc_i%2C%20c_j%7D_%7Bd%7D%3D%5Cxi%5E%7Bc_i%7D_%7Bd%7D%20%5Ccdot%20%5COmega%5E%7Bc_i%2Cc_j%7D_%7Bd%7D%20#0)

[where](https://www.codecogs.com/eqnedit.php?latex=%20%5Cgamma%5E%7Bc_i%2C%20c_j%7D_%7Bd%7D%3D%5Cxi%5E%7Bc_i%7D_%7Bd%7D%20%5Ccdot%20%5COmega%5E%7Bc_i%2Cc_j%7D_%7Bd%7D%20#0)
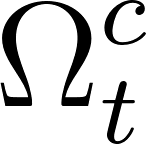
<https://www.codecogs.com/eqnedit.php?latex=%5COmega%5E%7Bc%7D_%7Bt%7D#0> [is the daily number of travelers from the UK to country *c*](https://www.codecogs.com/eqnedit.php?latex=%20%5Cgamma%5E%7Bc_i%2C%20c_j%7D_%7Bd%7D%3D%5Cxi%5E%7Bc_i%7D_%7Bd%7D%20%5Ccdot%20%5COmega%5E%7Bc_i%2Cc_j%7D_%7Bd%7D%20#0).

Assuming that the introduction of variant cases from the UK to each country *c* is essentially a non-homogeneous Poisson process (17–19), we estimate the probability of at least one introduction by time *t* (starting at time *t_0_*) using

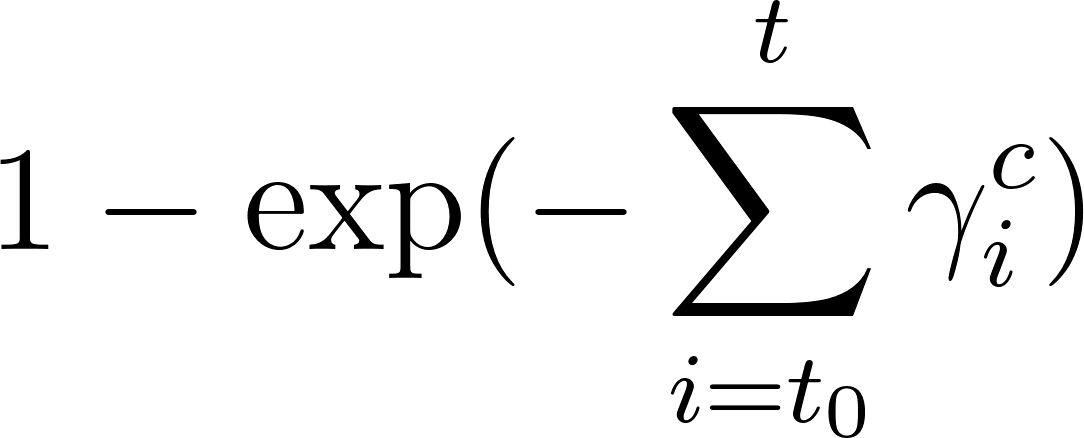
<https://www.codecogs.com/eqnedit.php?latex=1%20-%20%5Cexp(-%20%5Csum%5Et_%7Bi%3Dt_0%7D%20%5Cgamma%5E%7Bc%7D_%7Bi%7D)#0>.

***Estimating the prevalence of the variant in each country***

We assume the variant and wildtype share the same generation time (8). Let
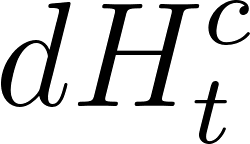
<https://www.codecogs.com/eqnedit.php?latex=dH%5E%7Bc%7D_t#0> denote the number of new COVID-19 hospitalizations in country *c* on day *t*. We assume that the number of new symptomatic and asymptomatic COVID-19 cases in country *c* on day *t* are given by

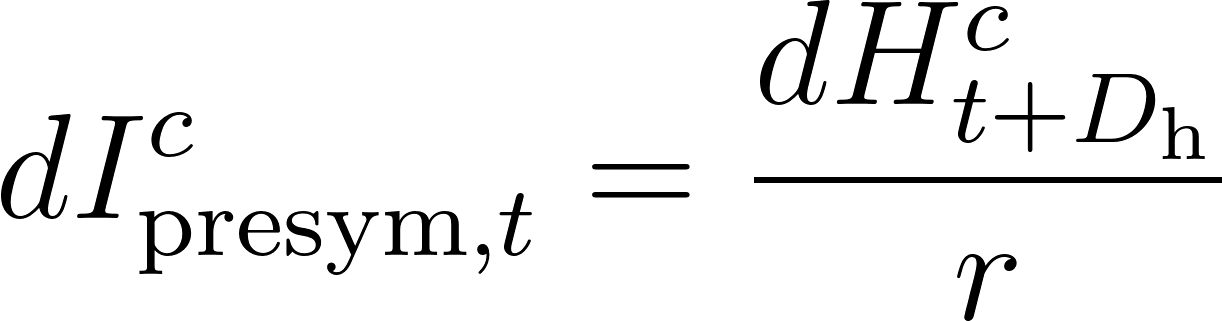
<https://www.codecogs.com/eqnedit.php?latex=dI%5E%7Bc%7D_%7B%5Ctext%7Bpresym%7D%2Ct%7D%20%3D%20%5Cfrac%7B%20dH%5E%7Bc%7D_%7Bt%2B%7BD_%7B%5Ctext%7Bh%7D%7D%7D%20%7D%20%7D%7Br%7D%20#0>

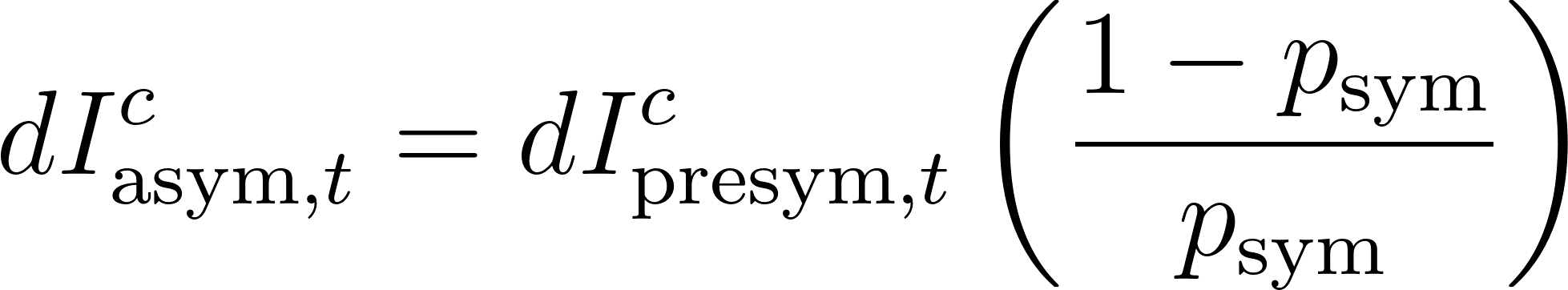
<https://www.codecogs.com/eqnedit.php?latex=dI%5E%7Bc%7D_%7B%5Ctext%7Basym%7D%2Ct%7D%20%3D%20dI%5E%7Bc%7D_%7B%5Ctext%7Bpresym%7D%2Ct%7D%5Cleft(%5Cfrac%7B1-p_%7B%5Ctext%7Bsym%7D%7D%7D%20%7Bp_%7B%5Ctext%7Bsym%7D%7D%7D%5Cright)%20#0>.

Let
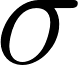
 [denote the ratio between the](https://www.codecogs.com/eqnedit.php?latex=%5Csigma#0) reproduction number of the 501Y variant and the reproduction number of the 501N variant. Then, the incidence of the 501Y and 501N variants are given by

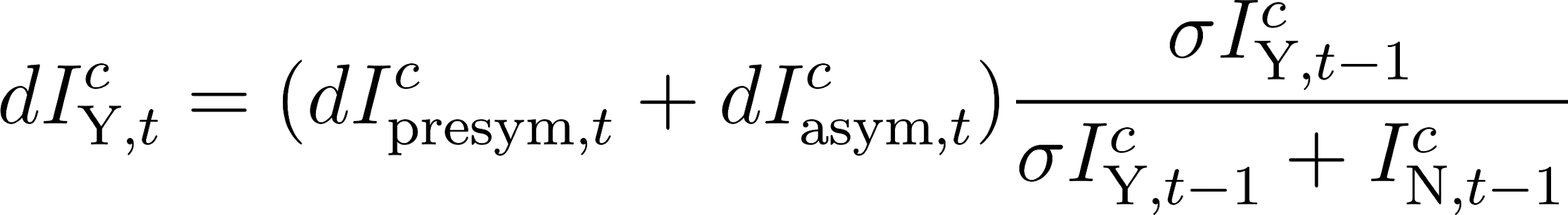
<https://www.codecogs.com/eqnedit.php?latex=dI%5E%7Bc%7D_%7B%5Ctext%7BY%7D%2Ct%7D%20%3D%20(dI%5E%7Bc%7D_%7B%5Ctext%7Bpresym%7D%2Ct%7D%20%2B%20dI%5E%7Bc%7D_%7B%5Ctext%7Basym%7D%2Ct%7D)%20%5Cfrac%7B%5Csigma%20I%5E%7Bc%7D_%7B%5Ctext%7BY%7D%2Ct-1%7D%7D%7B%5Csigma%20I%5E%7Bc%7D_%7B%5Ctext%7BY%7D%2Ct-1%7D%20%2B%20I%5E%7Bc%7D_%7B%5Ctext%7BN%7D%2Ct-1%7D%20%7D#0>

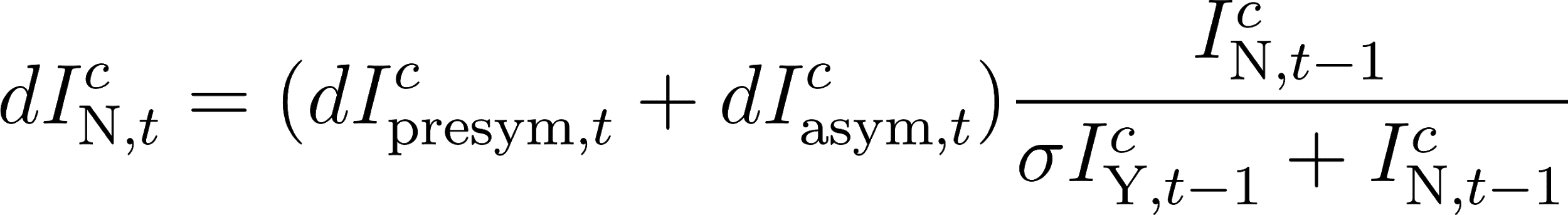
<https://www.codecogs.com/eqnedit.php?latex=dI%5E%7Bc%7D_%7B%5Ctext%7BN%7D%2Ct%7D%20%3D%20(dI%5E%7Bc%7D_%7B%5Ctext%7Bpresym%7D%2Ct%7D%20%2B%20dI%5E%7Bc%7D_%7B%5Ctext%7Basym%7D%2Ct%7D)%20%5Cfrac%7BI%5E%7Bc%7D_%7B%5Ctext%7BN%7D%2Ct-1%7D%7D%7B%5Csigma%20I%5E%7Bc%7D_%7B%5Ctext%7BY%7D%2Ct-1%7D%20%2B%20I%5E%7Bc%7D_%7B%5Ctext%7BN%7D%2Ct-1%7D%20%7D#0>.

We estimate the prevalence of the 501Y and 501N variants as a proportion of the population, as given by

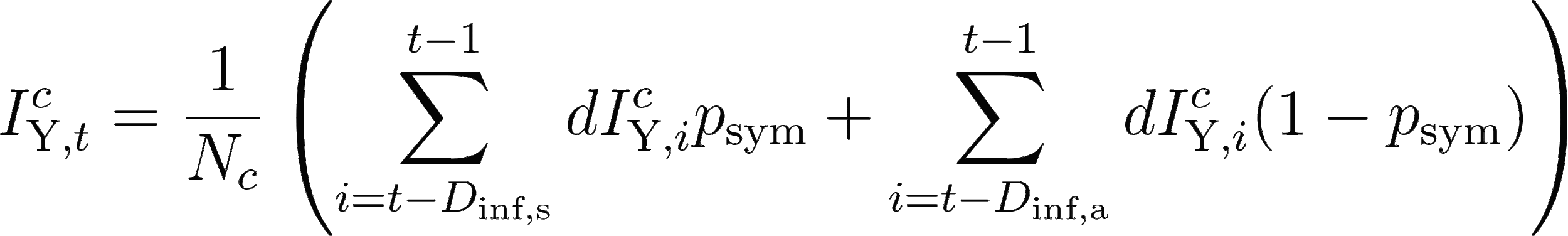
<https://latex-staging.easygenerator.com/eqneditor/editor.php?latex=I%5E%7Bc%7D_%7B%5Ctext%7BY%7D%2Ct%7D%20%3D%20%20%5Cfrac%7B1%7D%7BN_c%7D%5Cleft(%20%5Csum_%7Bi%20%3D%20t%20-%20D_%7B%5Ctext%7Binf%2Cs%7D%7D%7D%5E%7Bt-1%7D%20dI%5E%7Bc%7D_%7B%5Ctext%7BY%7D%2C%20i%7D%20p_%7B%5Ctext%7Bsym%7D%7D%20%20%2B%20%20%5Csum_%7Bi%20%3D%20t%20-%20D_%7B%5Ctext%7Binf%2Ca%7D%7D%7D%5E%7Bt-1%7D%20dI%5E%7Bc%7D_%7B%5Ctext%7BY%7D%2C%20i%7D(1-%20p_%7B%5Ctext%7Bsym%7D%7D)%5Cright)#0>

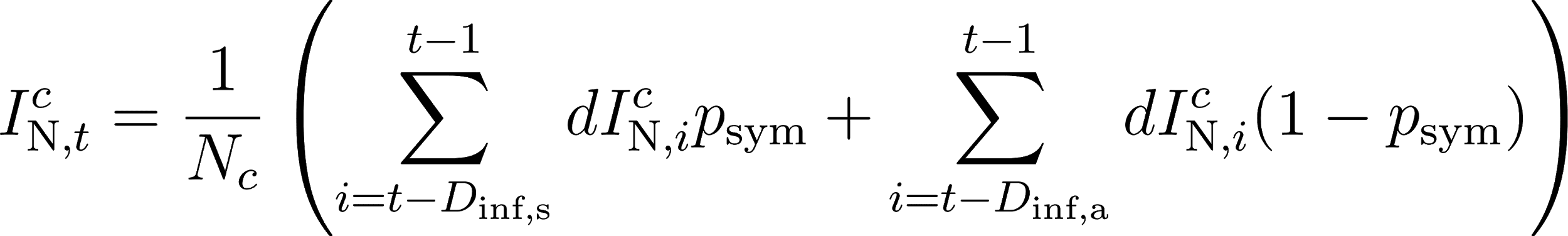
<https://www.codecogs.com/eqnedit.php?latex=I%5E%7Bc%7D_%7B%5Ctext%7BN%7D%2Ct%7D%20%3D%20%20%20%5Cfrac%7B1%7D%7BN_c%7D%5Cleft(%20%5Csum_%7Bi%20%3D%20t%20-%20D_%7B%5Ctext%7Binf%2Cs%7D%7D%7D%5E%7Bt-1%7D%20dI%5E%7Bc%7D_%7B%5Ctext%7BN%7D%2C%20i%7D%20p_%7B%5Ctext%7Bsym%7D%7D%20%20%20%2B%20%20%5Csum_%7Bi%20%3D%20t%20-%20D_%7B%5Ctext%7Binf%2Ca%7D%7D%7D%5E%7Bt-1%7D%20dI%5E%7Bc%7D_%7B%5Ctext%7BN%7D%2C%20i%7D(1-%20p_%7B%5Ctext%7Bsym%7D%7D)%5Cright)#0>.

At the start of each simulation, we assume that all cases on September 22, 2020 are the wildtype (501N). That is,
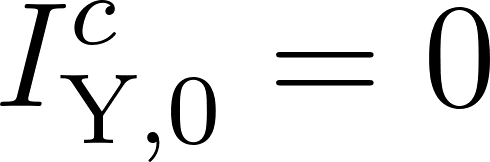
<https://www.codecogs.com/eqnedit.php?latex=I_%7B%5Ctext%7BY%7D%2C0%7D%5Ec%3D0#0> and
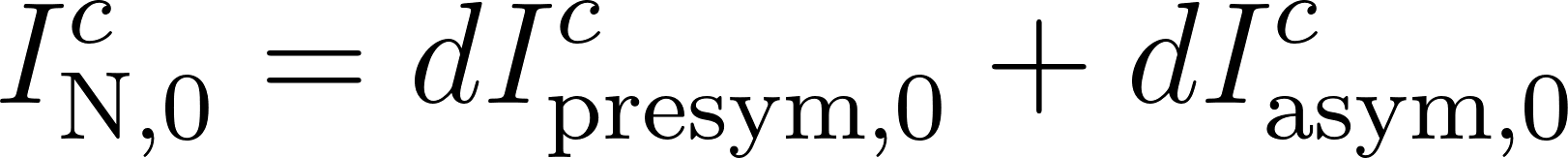
<https://www.codecogs.com/eqnedit.php?latex=I_%7B%5Ctext%7BN%7D%2C0%7D%5Ec%3DdI%5E%7Bc%7D_%7B%5Ctext%7Bpresym%7D%2C0%7D%20%2B%20dI%5E%7Bc%7D_%7B%5Ctext%7Basym%7D%2C0%7D%20#0>. Variant (501Y) cases are then introduced according to our travel model. Prior to the first introduction of the variant into country *c*, we draw a poisson random variable with a mean of
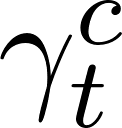
<https://latex-staging.easygenerator.com/eqneditor/editor.php?latex=%5Cgamma_t%5Ec#0> each day *t* (representing the number of asymptomatic and presymptomatic cases of 501Y traveling from the UK to *c*). At the first instance of a non-zero number of importations into country *c*, we set
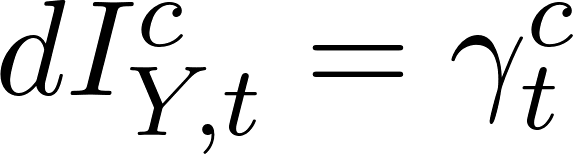
<https://www.codecogs.com/eqnedit.php?latex=dI_%7BY%2Ct%7D%5Ec%3D%5Cgamma_t%5Ec#0> and do not consider subsequent importations into *c*.

**Sensitivity Analyses**

Figures S1 and S2 provide a sensitivity analysis with respect to the relative transmission rate of the variant, with
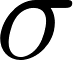
<https://www.codecogs.com/eqnedit.php?latex=%5Csigma#0> ranging from 25% to 75% more transmissible than the wildtype (Figures S1 and S2). Figure S3 provides a sensitivity analysis in which we simultaneously change the following two assumptions, both of which accelerate our estimates for importation times: (i) non-hospitalized symptomatic cases are allowed to travel and (ii) the mobility values are roughly doubled (scaled by 741/419) to account for the FaceBook data sample size (419 million regular users (1) out of a Europe population of 741 million (2) in 2020).

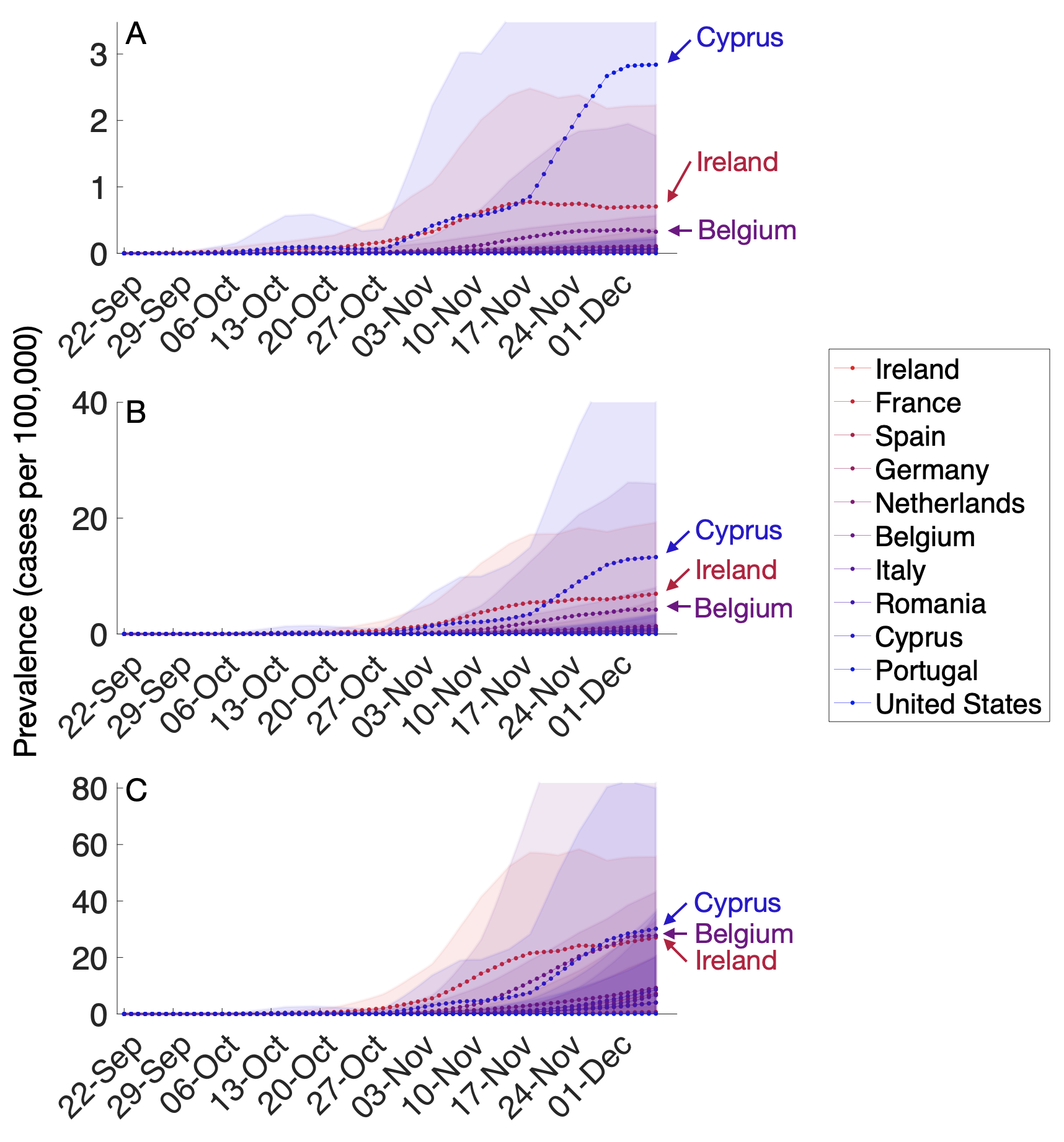

**Figure S1. Estimated daily prevalence of the 501Y variant of SARS-CoV-2 in 15 countries between September 22 and December 7, 2020, assuming the variant is**
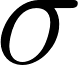
<https://www.codecogs.com/eqnedit.php?latex=%5Csigma#0> **= (A) 25%, (B) 50%, and (C) 75% more transmissible than 501N.** Points and bars indicate means and standard deviations based on 100 simulations. Countries with highest estimated prevalence on December 7th are labeled.

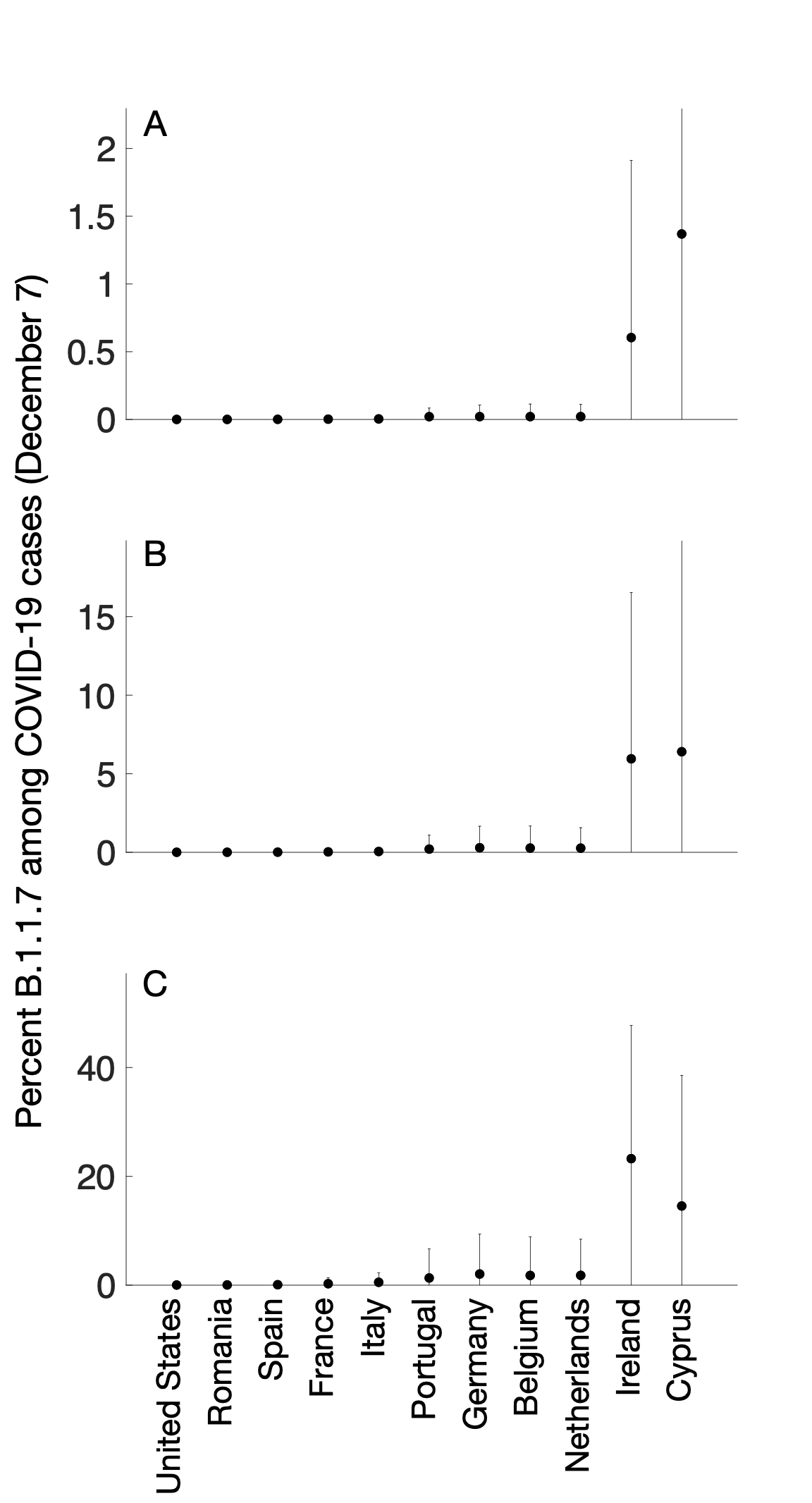

**Figure S2. Estimated proportion of the 501Y variant among all SARS-CoV-2 cases in 11 countries on December 7, 2020, assuming the variant is**
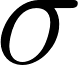
<https://www.codecogs.com/eqnedit.php?latex=%5Csigma#0> **= (A) 25%, (B) 50%, and (C) 75% more transmissible than 501N.** Points and bars indicate means and standard deviations across 100 simulations.

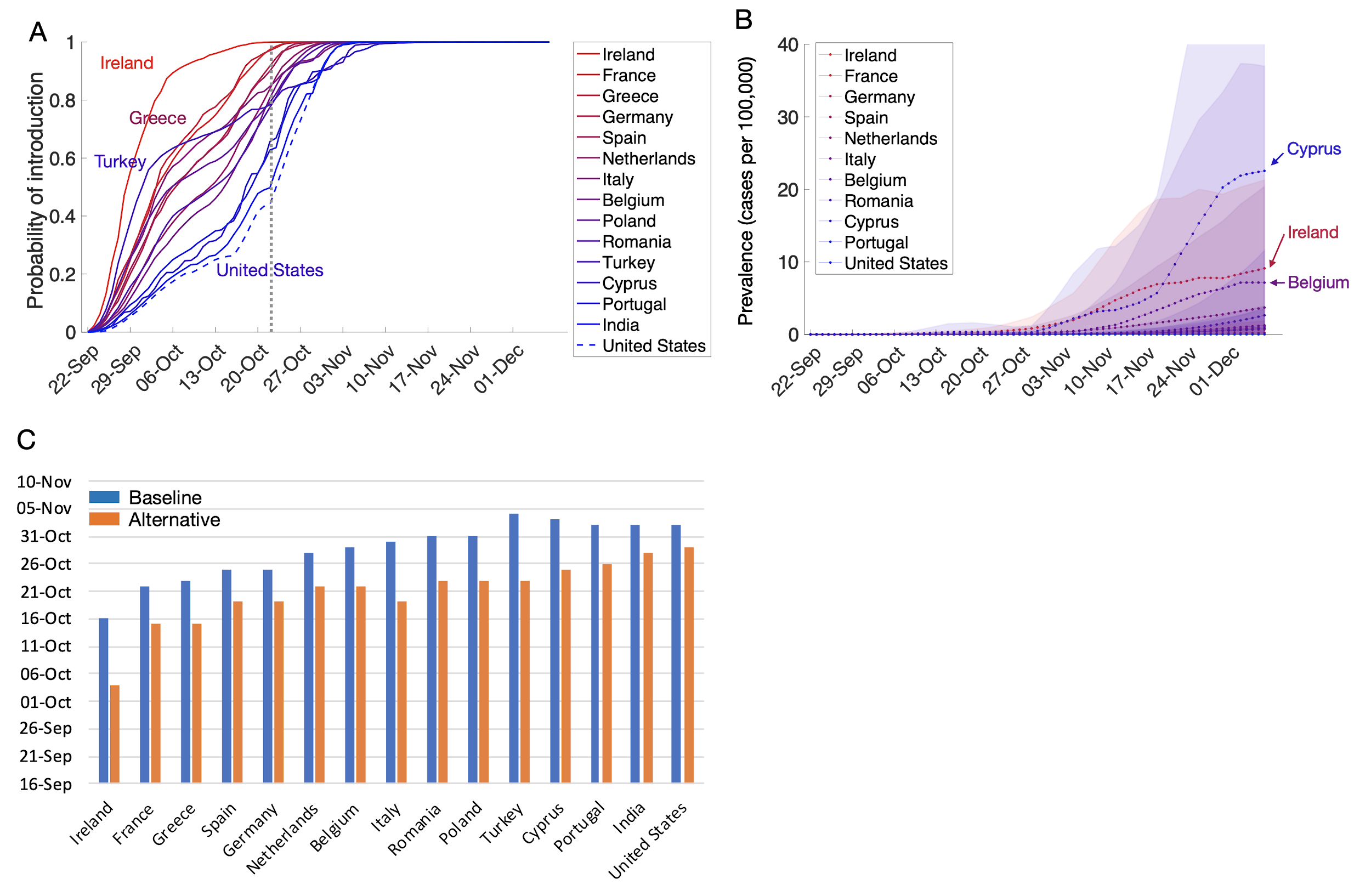

**Figure S3. Sensitivity analysis assuming that symptomatic cases can travel and increasing the mobility flows by a factor of 1.8.** (A) The probability that at least one person infected with the 501Y SARS-CoV-2 variant has arrived at the target country from the UK by the date indicated on the x-axis, based on Facebook mobility data. The dotted gray vertical line indicates October 23; line colors correspond to the relative risk of importations as of that date. (B) Estimated daily prevalence of the 501Y variant of SARS-CoV-2 in 11 countries between September 22 and December 7, 2020, assuming that the variant is
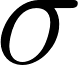
<https://www.codecogs.com/eqnedit.php?latex=%5Csigma#0>= 50% more transmissible than the 501N variant (16). Points and shaded bands indicate means and standard deviations based on 100 simulations. (C) The calendar day that the probability of a 501Y variant introduction first exceeds 0.8, for the baseline (blue) versus alternative (red) assumptions of elevated mobility and symptomatic travel.

**References**

1.	[Home - Facebook Data for Good [Internet]. [cited 2021 Jan 2]. Available from:](http://paperpile.com/b/m1AylW/WIMR6) <https://dataforgood.fb.com/>

2.	Gudbjartsson DF, Helgason A, Jonsson H, Magnusson OT, Melsted P, Norddahl GL, et al. Spread of SARS-CoV-2 in the Icelandic Population. N Engl J Med. 2020 Jun 11;382(24):2302–15.

3.	[King’s College London. No evidence of change in symptoms from new coronavirus variant [Internet]. King’s College London. 2021 [cited 2021 Mar 3]. Available from:](http://paperpile.com/b/m1AylW/xm7ou) <https://www.kcl.ac.uk/news/no-evidence-change-symptoms-new-coronavirus-variant>

4.	[Davies NG, Abbott S, Barnard RC, Jarvis CI, Kucharski AJ, Munday JD, et al. Estimated transmissibility and impact of SARS-CoV-2 lineage B.1.1.7 in England. Science [Internet]. 2021 Mar 3 [cited 2021 Mar 4]; Available from:](http://paperpile.com/b/m1AylW/ShGea) <https://science.sciencemag.org/content/early/2021/03/03/science.abg3055>

5.	[Official UK Coronavirus Dashboard [Internet]. [cited 2020 Dec 29]. Available from:](http://paperpile.com/b/m1AylW/9mOWr) <https://coronavirus.data.gov.uk/details/healthcare>

6.	[Coronavirus (COVID-19) Hospitalizations [Internet]. [cited 2021 Jan 3]. Available from:](http://paperpile.com/b/m1AylW/c8a8w) <https://ourworldindata.org/covid-hospitalizations>

7.	[CDC. Estimated Disease Burden of COVID-19 [Internet]. 2020 [cited 2020 Dec 28]. Available from:](http://paperpile.com/b/m1AylW/Ejghf) <https://www.cdc.gov/coronavirus/2019-ncov/cases-updates/burden.html>

8.	Leung K, Shum MHH, Leung GM, Lam TTY, Wu JT. Early transmissibility assessment of the N501Y mutant strains of SARS-CoV-2 in the United Kingdom, October to November 2020. Eurosurveillance. 2021 Jan 7;26(1):2002106.

9.	Backer JA, Klinkenberg D, Wallinga J. Incubation period of 2019 novel coronavirus (2019-nCoV) infections among travellers from Wuhan, China, 20–28 January 2020. Eurosurveillance. 2020 Feb 6;25(5):2000062.

10.	[Aleta A, Martín-Corral D, Pastore Y Piontti A, Ajelli M, Litvinova M, Chinazzi M, et al. Modelling the impact of testing, contact tracing and household quarantine on second waves of COVID-19. Nat Hum Behav [Internet]. 2020 Aug 5; Available from:](http://paperpile.com/b/m1AylW/pLxO) <http://dx.doi.org/10.1038/s41562-020-0931-9>

11.	[ISARIC. International Severe Acute Respiratory and Emerging Infections Consortium (ISARIC). COVID-19 Report: 19 [Internet]. 2020 [cited 2021 Feb 1]. Available from:](http://paperpile.com/b/m1AylW/atuLs) <https://media.tghn.org/medialibrary/2020/05/ISARIC_Data_Platform_COVID-19_Report_19MAY20.pdf>

12.	[Park N. Population estimates for the UK, England and Wales, Scotland and Northern Ireland - Office for National Statistics [Internet]. Office for National Statistics; 2020 [cited 2020 Dec 29]. Available from:](http://paperpile.com/b/m1AylW/bpSBa) <https://www.ons.gov.uk/peoplepopulationandcommunity/populationandmigration/populationestimates/bulletins/annualmidyearpopulationestimates/mid2019estimates>

13.	[World Population Prospects - Population Division - United Nations [Internet]. [cited 2021 Feb 3]. Available from:](http://paperpile.com/b/m1AylW/zzYQM) <https://population.un.org/wpp/Download/Standard/Population/>

14.	[Report 42 - Transmission of SARS-CoV-2 Lineage B.1.1.7 in England: insights from linking epidemiological and genetic data [Internet]. [cited 2021 Jan 3]. Available from:](http://paperpile.com/b/m1AylW/tcUeN) <https://www.imperial.ac.uk/mrc-global-infectious-disease-analysis/covid-19/report-42-sars-cov-2-variant/>

15.	[Leung K, Shum MHM, Leung GM, Lam TTY, Wu JT. Early empirical assessment of the N501Y mutant strains of SARS-CoV-2 in the United Kingdom, October to November 2020 [Internet]. bioRxiv. medRxiv; 2020. Available from:](http://paperpile.com/b/m1AylW/mWJGg) <http://medrxiv.org/lookup/doi/10.1101/2020.12.20.20248581>

16.	[Davies NG, Barnard RC, Jarvis CI, Kucharski AJ. Estimated transmissibility and severity of novel SARS-CoV-2 Variant of Concern 202012/01 in England. medRxiv [Internet]. 2020; Available from:](http://paperpile.com/b/m1AylW/ZUHHk) <https://www.medrxiv.org/content/10.1101/2020.12.24.20248822v1.full-text>

17.	Wang L, Wu JT. Characterizing the dynamics underlying global spread of epidemics. Nat Commun. 2018 Jan 15;9(1):218.

18.	Scalia Tomba G, Wallinga J. A simple explanation for the low impact of border control as a countermeasure to the spread of an infectious disease. Math Biosci. 2008 Jul;214(1-2):70–2.

19.	Wu JT, Leung K, Leung GM. Nowcasting and forecasting the potential domestic and international spread of the 2019-nCoV outbreak originating in Wuhan, China: a modelling study. Lancet. 2020 Feb 29;395(10225):689–97.

20.	[Facebook: Europe monthly active users by quarter 2020 [Internet]. [cited 2021 Mar 6]. Available from:](http://paperpile.com/b/m1AylW/v4laY) <https://www.statista.com/statistics/745400/facebook-europe-mau-by-quarter/>

21.	[European Union (EU) - total population 2020 [Internet]. [cited 2021 Mar 6]. Available from:](http://paperpile.com/b/m1AylW/SG5r4) <https://www.statista.com/statistics/253372/total-population-of-the-european-union-eu/>
